# A Modified Mediterranean Ketogenic Diet mitigates modifiable risk factors of Alzheimer’s Disease: a serum and CSF-based metabolic analysis

**DOI:** 10.1101/2023.11.27.23298990

**Authors:** Annalise Schweickart, Richa Batra, Bryan J. Neth, Cameron Martino, Liat Shenhav, Anru R. Zhang, Pixu Shi, Naama Karu, Kevin Huynh, Peter J. Meikle, Leyla Schimmel, Amanda Hazel Dilmore, Kaj Blennow, Henrik Zetterberg, Colette Blach, Pieter C Dorrestein, Rob Knight, Alzheimer’s Gut Microbiome Project Consortium, Suzanne Craft, Rima Kaddurah-Daouk, Jan Krumsiek

## Abstract

Alzheimer’s disease (AD) is influenced by a variety of modifiable risk factors, including a person’s dietary habits. While the ketogenic diet (KD) holds promise in reducing metabolic risks and potentially affecting AD progression, only a few studies have explored KD’s metabolic impact, especially on blood and cerebrospinal fluid (CSF). Our study involved participants at risk for AD, either cognitively normal or with mild cognitive impairment. The participants consumed both a modified Mediterranean-ketogenic diet (MMKD) and the American Heart Association diet (AHAD) for 6 weeks each, separated by a 6-week washout period. We employed nuclear magnetic resonance (NMR)-based metabolomics to profile serum and CSF and metagenomics profiling on fecal samples. While the AHAD induced no notable metabolic changes, MMKD led to significant alterations in both serum and CSF. These changes included improved modifiable risk factors, like increased HDL-C and reduced BMI, reversed serum metabolic disturbances linked to AD such as a microbiome-mediated increase in valine levels, and a reduction in systemic inflammation. Additionally, the MMKD was linked to increased amino acid levels in the CSF, a breakdown of branched-chain amino acids (BCAAs), and decreased valine levels. Importantly, we observed a strong correlation between metabolic changes in the CSF and serum, suggesting a systemic regulation of metabolism. Our findings highlight that MMKD can improve AD-related risk factors, reverse some metabolic disturbances associated with AD, and align metabolic changes across the blood-CSF barrier.

## Introduction

Alzheimer’s disease (AD) is the fifth-leading cause of death in Americans over the age of 65^1^. Despite recent advancements with approval of novel therapeutics, there remains no cure or established preventive therapy for AD^2–5^. While age, genetics, and family history are prominent risk factors for the development of AD^6,7^, environmental and lifestyle factors significantly influence onset, progression, and severity of the disease^8^. Modifiable conditions such as hypertension^9^, type 2 diabetes mellitus^10^, and dyslipidemia^11^ have been identified as risk factors for AD, highlighting the potential for protective measures before the clinical onset of symptoms. Dietary and related lifestyle interventions have emerged over the last decades as a way to mitigate such modifiable metabolic risk factors^12^.

One dietary approach that has gained recent attention is the ketogenic diet (KD), a high-fat, low-carbohydrate diet that has been successfully used to treat drug-resistant epilepsy^13,14^ and showed promising results in cancer therapy^15,16^. In KD, the metabolic system is rewired to use fat as the primary source of energy instead of carbohydrates. Catabolism of fat via lipolysis leads to the production of ketone bodies, which replaces glucose as the body’s main fuel source^15^. Consequently, KD induces various system-wide metabolic alterations which lead to positive health benefits, including increased insulin sensitivity^17^, weight reduction in overweight individuals, and improved lipid profiles^18^. KD has also been linked to neuroprotective effects, including the promotion of more efficient mitochondrial activity and reduction of mitochondria-derived reactive oxygen species^19^, as well as a decrease in neuronal excitotoxicity^20,21^. Given the role of metabolic dysregulation, oxidative stress, and excitotoxicity in the pathogenesis of AD^13^, the potential metabolic shifts related to KD could provide beneficial alterations in disease progression^19,22^. However, there have been only a few KD studies in humans that characterize its metabolic effects, and none employed metabolic profiling in blood and CSF simultaneously^23,24^. As such, the potential impact of a KD on the systemic and central nervous system-specific metabolic dysregulation of AD remains uncertain.

In this study, we characterized the peripheral and central metabolic effects of dietary interventions in participants at risk for AD. We used samples from a randomized crossover pilot trial of a Modified Mediterranean-Ketogenic Diet (MMKD) or the American Heart Association Diet (AHAD) (**Figure 1**) in a cohort of prediabetic participants that were either mildly cognitively impaired (MCI, n = 9) or cognitively normal (CN, n = 10)^20^. In a series of studies using the same cohort, we previously found that MMKD leads to increased Aβ42 and decreased tau in CSF^20^, changes in the gut microbiome^21^, and reversal of serum-based AD-associated lipid signature^25^. In the current study, we broaden the scope of blood biomarkers by using an NMR-metabolomics platform that measured amino acids, ketone bodies, triglycerides, and lipoprotein levels and lipoprotein composition. Moreover, this is the first exploration of diet-related metabolic changes in CSF. In both serum and CSF, we determine the MMKD-associated molecular changes, discuss their association to AD pathology, and outline their potential to counteract modifiable risk factors.

**Figure 1:**
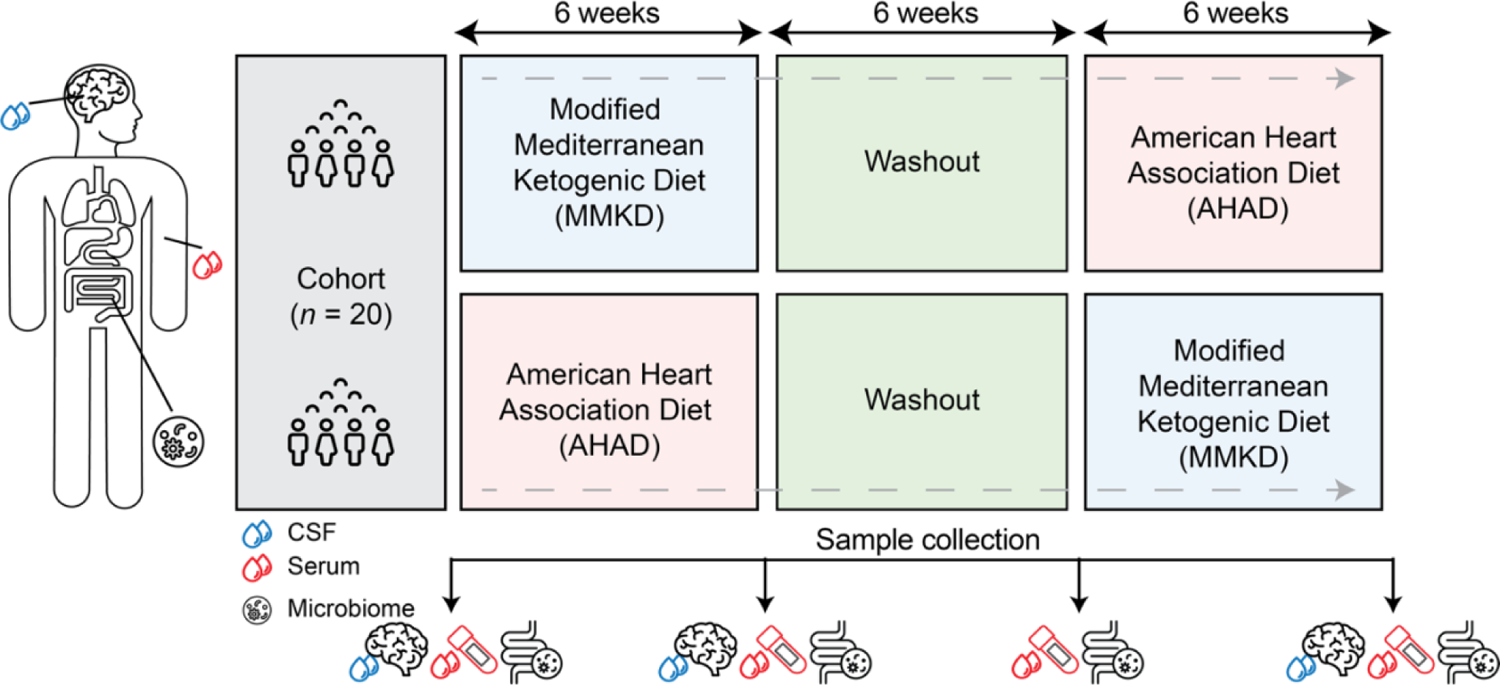
Cross-over design of the diet study: In the study, 20 participants were enrolled. They were randomized into two groups: one followed the modified Mediterranean ketogenic diet (MMKD) and the other followed the American Heart Association diet (AHAD). Each group adhered to their respective diet for 6 weeks, followed by a 6-week washout period, after which they switched to the other diet. Throughout the study, samples from cerebrospinal fluid (CSF), serum, and microbiome were collected from each participant at various time points.

## Results

### MMKD alters serum levels of amino acids, inflammatory markers, and lipoproteins

For both MMKD and AHAD, linear mixed-effects models were used to assess diet-induced changes, with metabolites as outcomes, the diet timepoint (pre/post), cognitive status, and a diet-cognition interaction term as predictors, and a subject-based random effect. The interaction term was included to examine whether there was a relationship between the effect of diet and cognitive status on a metabolite’s abundance; for example, if the direction or magnitude of a metabolite’s change due to the ketogenic diet was affected by a subject’s cognitive status. For the AHAD, no metabolites were found to be significantly changed from pre- to post timepoints in serum or CSF (**Supplementary Tables 2 and 4**) and will thus not be further discussed.

In the MMKD, we found 38 metabolites that associated with diet and 92 metabolites that associated with cognitive status (false discovery rate, FDR<20%; see Figure 2a**, Supplementary Table 1**). Glycine was the only metabolite associated with both diet and cognitive status, and no metabolites showed an interaction between the two factors (**Supplementary Table 1**). Of the metabolites whose abundance was significantly affected by the MMKD, we focused on changes with a potential relevance to metabolic impairment in AD:

**Figure 2:**
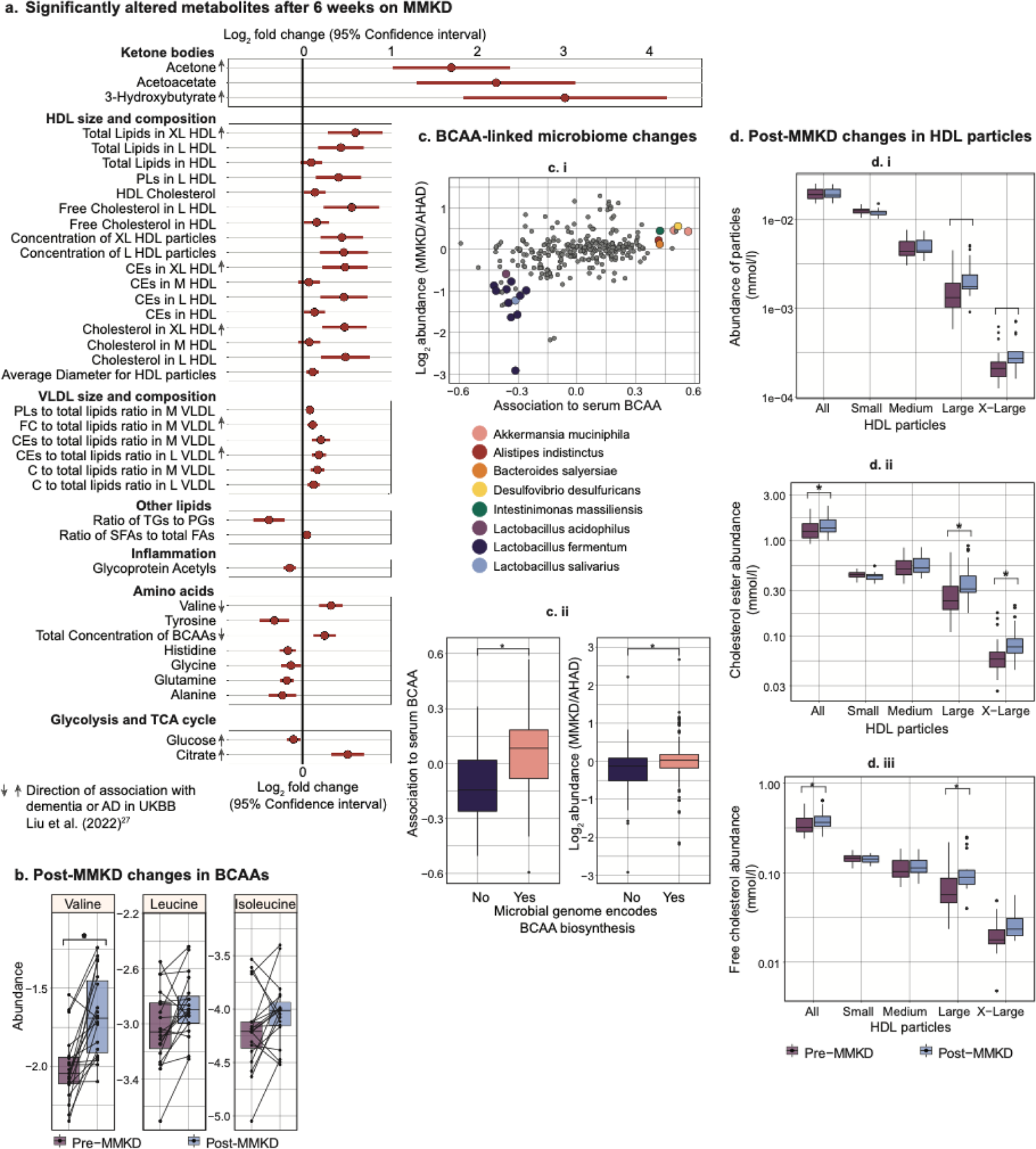
Serum metabolite associated with the modified Mediterranean ketogenic diet (MMKD). **a.** Fold change of significantly altered metabolites after 6 weeks on MMKD (n = 19). The arrows next to the metabolite names indicate the effect direction of these metabolites in dementia or AD based on the UKBB cohort^27^. b. Changes in the levels of the three branched-chain amino acids (BCAAs) pre- and post-diet. Each line represents the trend in a single individual. c. i) Microbiome species that show differential abundance by diet and are significantly correlated with serum BCAA (p = 3.9e-13). ii) Species whose genome encodes BCAA biosynthesis have a significantly higher association to serum BCAA and are significantly more enriched post-MMKD as compared to AHAD. d. Pre- and post-diet changes in total HDL and size-specific HDL particle concentrations, cholesterol ester, and free cholesterol content (mmol/l). Abbreviations: HDL = high-density lipoprotein, PLs = phospholipids, CEs = cholesteryl esters, VLDL = very low-density lipoprotein, FC = free cholesterol, C = cholesterol, TGs = triglycerides, PGs= phosphoglycerides, SFAs = saturated fatty acids, FA = fatty acids, BCAAs = branched-chain amino acids.

### Ketone bodies

We detected an increase in serum ketone bodies namely acetone (Log_2_ fold change, LFC = 1.69, 95 % confidence interval, CI = 1.02 to 2.36, adjusted p-value, adj.p = 1.5e-2), acetoacetate (LFC = 2.20, adj.p = 1.2e-2), and 3-hydroxybutyrate (LFC = 2.98, adj.p = 7e-3), for individuals consuming the ketogenic diet, while serum glucose levels decreased (LFC = −0.10, 95% CI = −0.18 to −0.02, adj. p = 1.7e-1, Figure 2a). This aligns with the expected impact of a ketogenic diet, which is known to shift the body’s metabolism towards utilization of ketone bodies as an energy source instead of glucose^26^. Remarkably, previous studies using the UK Biobank cohort (UKBB) have shown a positive association of plasma ketone bodies with Alzheimer’s disease incidence^27^. While the mechanism behind this relationship is unclear, beyond the context of a ketogenic diet, ketone bodies are produced as a response to insulin resistance in type 2 diabetes, and conditions of oxidative stress and hypoxia^28^. Since people with type 2 diabetes have an increased incidence of AD, the positive association between ketone bodies and AD in UKBB could be mediated by the prevalence of type 2 diabetes^29^.

### Amino acids

Branched-chain amino acids (BCAAs), which include valine, leucine, and isoleucine, are essential amino acids and are primarily obtained through dietary intake^30^. We observed an increase in the abundance of valine after the ketogenic diet (LFC = 0.33, adj. p = 3e-2), while leucine and isoleucine levels remained unaffected (Figure 2b). Previous research in limited cohorts has indicated that plasma BCAA levels tend to increase during fasting and short periods of starvation^31,32^. Both fasting and the ketogenic diet lead to ketogenesis, potentially related to elevated BCAA levels^33^. However, the reason for the observed increase in only valine levels within the BCAAs remains unclear. BCAAs have drawn particular attention as a decrease in their abundance in the blood has been linked to cognitive decline^34^, progression of Parkinson’s disease (PD)^35^, and incident dementia^27^. Our results suggest that a ketogenic diet may help rectify this imbalance by raising valine levels.

Notably, in addition to diet, the gut microbiome also contributes to BCAA biosynthesis^36^, and in PD, changes in the microbiome composition have been shown to be associated with BCAA levels^35^. In our study, post-MMKD, there was an increase in the abundance of gut microbiome species positively associated with serum BCAA compared to AHAD. Furthermore, species with genomes that encode for BCAA biosynthesis were notably more enriched after the keto diet (Figure 2c). These findings implicate diet-directed gut microbiome modification that provides an alternative source of BCAA.

### GlycA

Serum glycoprotein acetylation (GlycA) levels were decreased after following a ketogenic diet (LFC = −0.14, adj.p = 2.1e-2). GlycA is an indicator of systemic inflammation as it measures the glycosylation of acute phase reactants proteins which the liver releases in response to pro-inflammatory cytokines^37,38^. GlycA levels have been associated with worse global cognitive performance in a population of middle-aged and older adults (n = 15,105)^39^, which aligns with other findings of an overall association between inflammation and cognitive impairment^40,41^. The reduction of GlycA due to MMKD may indicate the diet’s anti-inflammatory effects, which may provide neuroprotection^42,43^.

### Lipoprotein size and lipid composition

The total concentration of HDL particles did not significantly change due to MMKD, but the average HDL particle size significantly increased, corresponding with increased concentrations of large and extra-large HDL particles (L-HDL: LFC = 0.48, adj. p = 1.8e-1, XL-HDL: LFC = 0.42, adj. p = 1.8e-1, Figure 2d). These particle size changes are accompanied by a significant increase in total HDL cholesterol (HDL-C: LFC = 0.15, adj. p = 1.8e-1), indicating that the increase in size of these particles may have been due to an increase in their cholesterol content. HDL particles are integral players in the reverse cholesterol transport (RCT) pathway, as they deliver excess cholesterol stores from peripheral tissues to the liver via the plasma, which is then excreted through the bile^44^. Midlife HDL-C levels have been found to be inversely associated with late-life MCI and dementia, indicating that an increase in HDL-C, as seen with the MMKD, could be protective against cognitive decline^45^.

### MMKD changes levels of amino acids in CSF

The Nightingale platform measured 28 metabolites in the CSF. 18 out of these 28 metabolites were significantly changed (FDR < 5%) after MMKD (Figure 3), but no metabolites were different due to cognitive status, nor any significant metabolic interactions between the two (Supplementary Table 3). Comparing the MMKD-affected metabolites in CSF to their counterparts in serum where applicable, isoleucine and glutamine were the only metabolites that significantly changed in the same direction due to the diet (Figure 3a).

**Figure 3:**
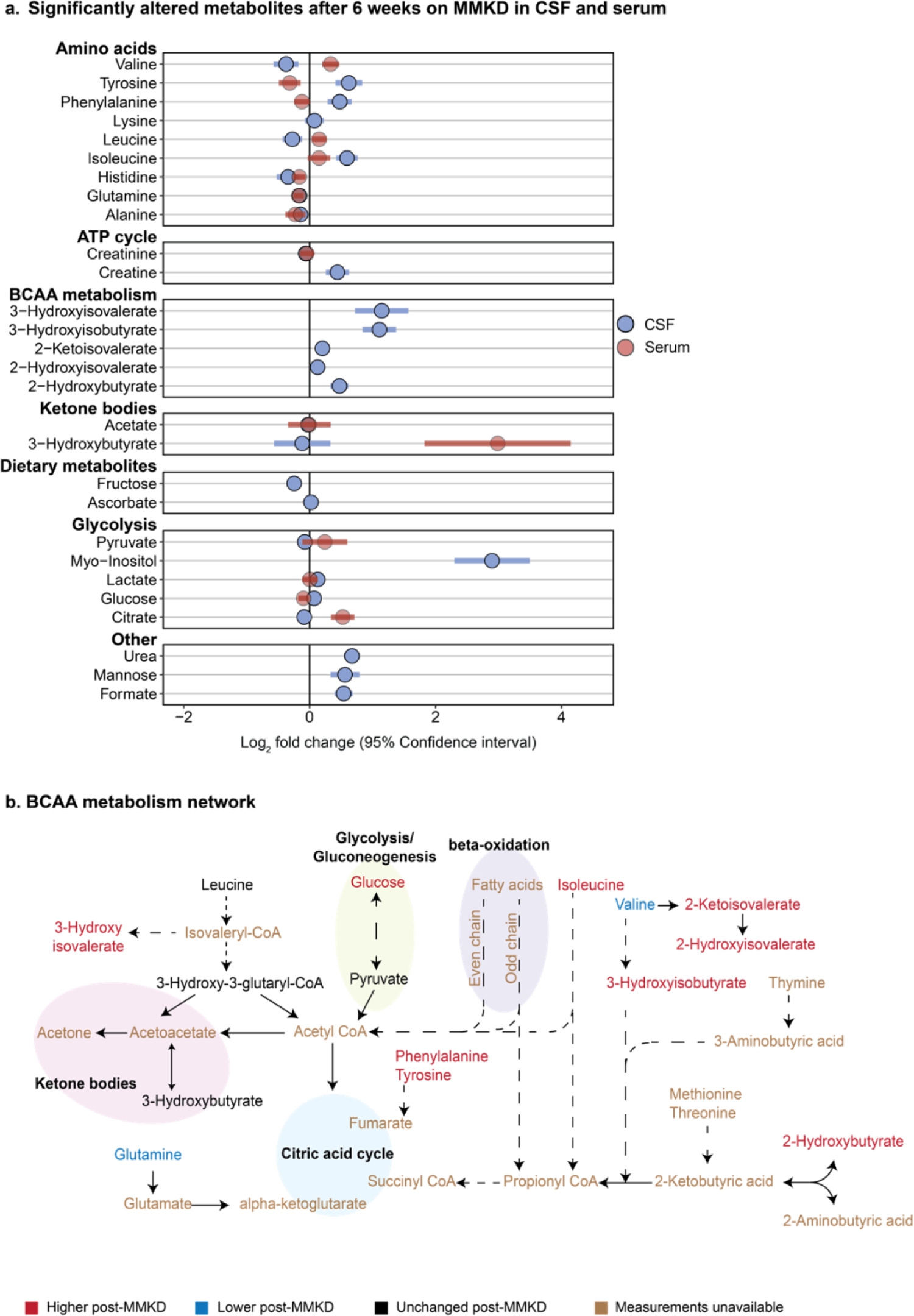
CSF metabolites associated with the modified Mediterranean ketogenic diet (MMKD). **a.** Fold change of significantly altered metabolites after 6 weeks on MMKD (n = 19) with 95% confidence interval. Light blue circles represent the metabolites altered in CSF. The corresponding change of each metabolite measured in serum is shown in red. **b.** BCAA metabolism network with MMKD-altered metabolites colored to reflect effect direction. Blue represents a decrease post-diet, red represents an increase post-diet, black represents unchanged, and brown represents unavailable measurements. Dotted lines indicate the presence of intermediate steps between two metabolites, while solid lines are a direct conversion.

#### Amino acids

Amino acids that were increased in the CSF after consumption of the MMKD were tyrosine (LFC = 0.62, adj. p = 1.8e-5), phenylalanine(LFC = 0.48, adj. p = 2.2e-4), and isoleucine (LFC = 0.59, adj. p = 1.1e-5), while glutamine (LFC = −0.16, adj. p = 2.6e-3) and valine (LFC = −0.37, adj. p = 4.8e-2) were decreased. This pattern of changes in CSF amino acids shows the opposite effect of the MMKD seen in serum, where we observed a systemic decrease in amino acids and an increase in BCAAs. Our previous metabolomic analysis of postmortem brain tissue has shown a positive correlation of BCAAs with AD traits^46^.

#### BCAA degradation

Following consumption of the MMKD, there was a marked increase in CSF concentrations of BCAA degradation products^47,48^ including 3-hydroxyisobutyrate (LFC = 1.109, adj. p = 3.9e-7), 2-hydroxyisovalerate (LFC = 0.13, adj. p = 2.6e-3), 2-ketoisovalerate (LFC = 0.2, adj. p = 9.7e-4), and 3-hydroxyisovalerate (LFC = 1.1, adj. p = 2.3e-4). This, along with the decreased concentrations of valine in CSF following MMKD, suggests an increased BCAA catabolism (Figure 3b). BCAA degradation plays a major role in generating energy substrates for the tricarboxylic acid (TCA) cycle, indicating that an MMKD-induced increase in BCAA degradation could lead to altered energetic pathways in CSF^49^.

To assess the role of the identified MMKD-altered CSF metabolites in relation to the pathology of AD and dementia, an extensive literature review was performed. The few existing studies examining AD-associated metabolic dysregulation in CSF show limited coverage and inconclusive if not conflicting results^50–53^. Using valine as an example, Ibáñez et al.^53^ used capillary-electrophoresis mass spectrometry on the CSF of 85 subjects and found an increase of valine in the CSF in subjects diagnosed with AD, while Berezhnoy et al.^54^ found a decrease in CSF valine in AD patients after performing NMR metabolomics on the CSF of 71 patients. It is therefore unclear how the observed alterations in CSF induced by an MMKD may affect AD disease development or progression.

### MMKD-induced metabolic changes show cross-compartmental relationships

After establishing the metabolic signature of the MMKD in serum and CSF, we investigated the crosstalk between the two. The blood-CSF barrier is required to maintain the composition of the CSF, thereby sustaining the homeostasis necessary for the proper functioning of the brain^55^. Consequently, only selected molecules essential to brain function are able to cross this barrier. Given this tightly regulated barrier, a one-to-one correspondence of metabolites across the compartments is unlikely. However, we expect a systemic coregulation of central and peripheral metabolism.

To assess this, we performed a correlation network analysis on the magnitude of changes between all metabolites affected by the MMKD, defined as the difference between metabolite’s pre- and post-diet abundance. This analysis models the functional relationship between two cross-compartmental metabolites by capturing how similarly metabolites change due to the MMKD. There were 43 significant correlations (FDR < 5%), 20 of which were between CSF metabolites and serum ketone bodies (Figure 4). Of note, there was no correlation in changes between serum ketone bodies and CSF ketone bodies. Specifically, in contrast to serum measurements, 3-hydroxybutyrate levels in the CSF did not show significant alterations post-MMKD. A potential reason for this discrepancy could be the effective utilization of ketone bodies in the brain, requiring the conversion of 3-hydroxybutyrate to acetoacetate and thus potentially stabilizing the CSF levels of acetoacetate^56^. In addition, compared to 3-hydroxybutyrate, acetoacetate is estimated to have double the transport rate through the BBB via MCT (monocarboxylate transporter), exhibiting competitive inhibition of the transport of 3-hydroxybutyrate^56–58^. This is further supported by a previously reported PET-based analysis on this cohort which showed an increase in one of the ketone bodies, acetoacetate, in the brain following MMKD^20^.

**Figure 4:**
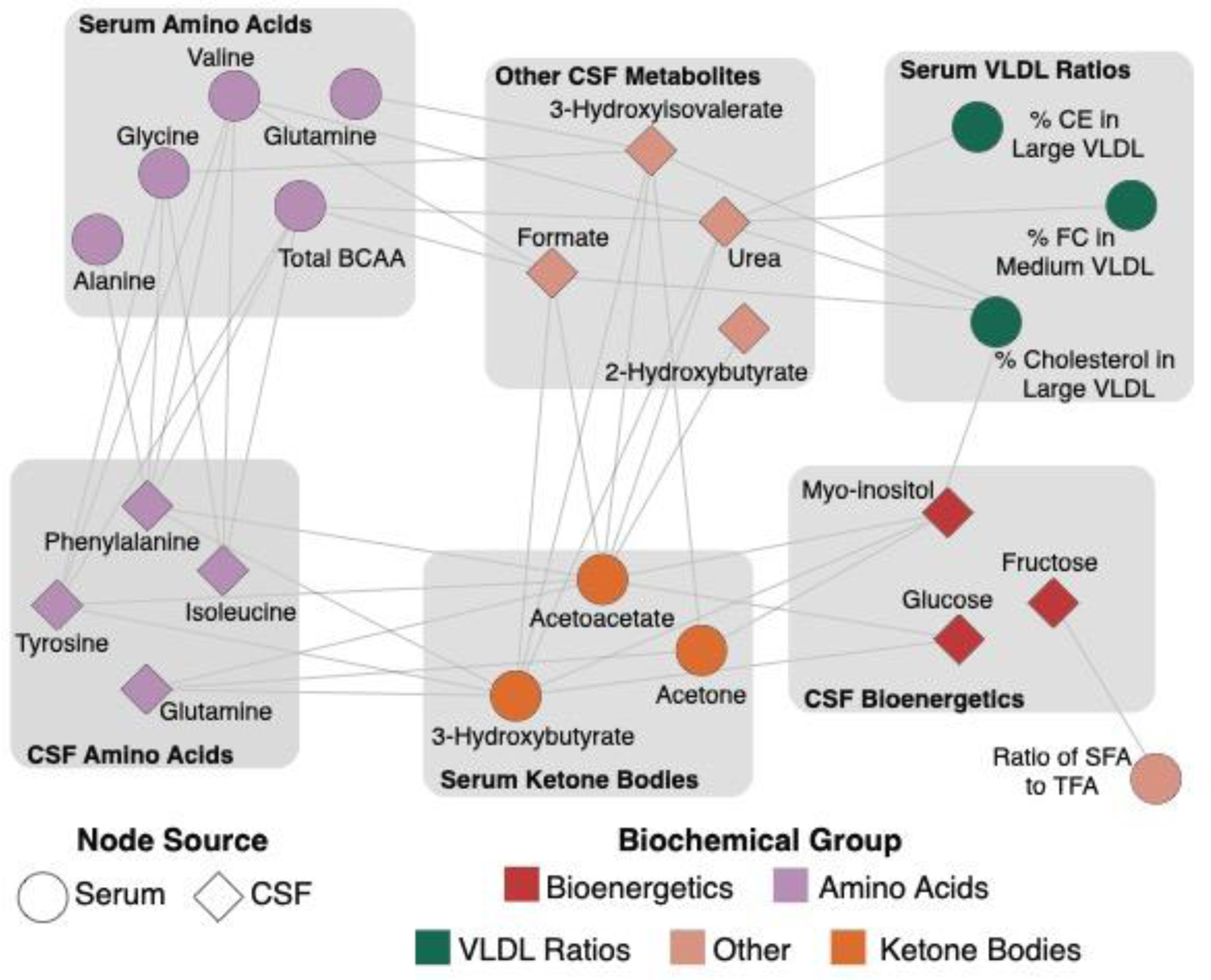
Bipartite graph between CSF and serum metabolites. Each node represents a metabolite. Ellipses depict metabolites measured in serum, and diamonds represent metabolites measured in CSF. Each edge represents a significant (FDR < 5%) correlation of the magnitude of changes pre- to post-diet between two metabolites. Node color represents their biochemical groups.

Overall, these relationships between serum ketone bodies and CSF metabolites confirm our hypothesis that the ketogenic diet leads to concurrent changes in metabolites across the blood-CSF barrier.

## Discussion

Modifiable risk factors, such as dietary habits, play a substantial role in the onset and progression of Alzheimer’s disease (AD)^59^. Though the ketogenic diet (KD) appears promising in diminishing metabolic threats and possibly influencing AD’s course^60^, only a few studies have delved into KD’s metabolic effects on blood and cerebrospinal fluid (CSF). In this study, we report the metabolic impact of dietary interventions simultaneously in serum and CSF of patients at risk for AD. Our study is based on 20 prediabetic participants who were on MMKD and AHAD for 6 weeks each with 6 weeks washout period between the two diets. We performed metabolic profiling of their serum and CSF samples, as well as metagenomic profiling of their fecal samples. Our findings show that the MMKD had a widespread metabolic impact on both serum and CSF, while the AHAD had minimal impact. Interestingly, the metabolic impact of the MMKD was independent of the cognitive status of the individuals. In the following, we discuss the key findings of our study indicating the positive impact of MMKD.

The changes in serum due to the MMKD show the diet’s potential for AD metabolic signature reversal. Our findings show that the MMKD reduces the inflammatory marker GlycA, indicating a decrease in systemic inflammation due to the diet. Systemic inflammatory markers have been shown to have an inverse relationship with cognitive abilities^39–41,61^. Furthermore, MMKD altered the BCAA-associated gut microbiome and increased circulating valine levels. Blood valine levels have been shown to be inversely associated with cognitive performance (ADAS-Cog13)^62^. These metabolic changes suggest that the MMKD could provide a therapeutic advantage by reversing the levels of the markers tied to the disease signature.

MMKD has the potential to mitigate modifiable risk factors of AD including those associated with cardiovascular disease and elevated body mass index (BMI). One of the key characteristics of a high-fat low-carb diet is its effect on the lipid profile^63^. In our study, an increase in HDL-C levels was observed post-MMKD. High HDL-C is conducive to improved cardiovascular health, a modifiable risk factor of AD^64^. Moreover, mid-life levels of HDL cholesterol show a consistent negative association with the development of cognitive impairment later in life^45^. In addition, previous work on this cohort reported that MMKD led to a significant decrease in weight (p < 1e-6)^20^. Overweight and obese body mass index (BMI) in mid-life is associated with an increased risk of dementia compared with normal BMI^65^. Taken together, MMKD which simultaneously increases HDL cholesterol, reduces systemic inflammation, and decreases weight could provide protection against developing AD later in life^66^.

Post-MMKD alterations in the metabolic profile of the CSF point to an increase in amino acid levels besides glutamine and valine, which decreased; the decrease in valine was accompanied by an increase in BCAA degradation metabolites, indicating a potential upregulation of BCAA catabolism. There is limited research in linking cognitive performance to BCAAs and their metabolism in CSF and thus it is unclear if the dysregulation of BCAA metabolism in CSF due to the MMKD would have any disease-modifying effect. However, we have previously shown that the CSF-based AD-biomarker profile of these individuals improves post-MMKD, as indicated by an increase in Aβ42 levels (in both MCI and CN) and a decrease in tau (only in MCI)^20^. Therefore, while prior research was unable to link the observed metabolic changes with AD pathology, it is likely that post-MMKD metabolic changes in CSF contribute to the observed reversal of AD-related pathological changes observed in this cohort.

Furthermore, when examining the changes induced by MMKD in both serum and CSF, we observed that the diet-related metabolic alterations were linked across the blood-CSF barrier. The substantial metabolic effects of the MMKD in CSF and the strong relationship between ketone body changes in serum and CSF metabolites indicate the potential of this dietary intervention to alter brain metabolism, which is dysregulated in AD^67^. We and others have shown that ketogenic interventions increase ketone body uptake in the brain^20,68,69^. With increased ketone bodies as an energy substitute for glucose in the serum, a similar increase in the brain could be beneficial, to compensate for the cerebral glucose hypometabolism in AD.

The main limitation of our study is the small sample size, which restricts the generalizability of our results. It also limits our ability to incorporate potential additional covariates in statistical models, such as the *APOE4* genotype, a known genetic component of Alzheimer’s disease^70^. The fact that all participants were prediabetic limits the generalizability of our findings to metabolically healthy individuals although provides a basis to assess metabolic response in those with impaired systemic metabolism. Another limitation of our study is the poor metabolic coverage in CSF due to the platform used, which only included 28 biomarkers. This limits our ability to obtain a full picture of the impacts of the ketogenic diet on the CSF.

Taken together, our study highlights the positive impact of MMKD on metabolic health in the context of AD. We showed that AD-related modifiable risk factors can be improved, AD-linked metabolic alterations can be partially reversed, and importantly that diet-related metabolic changes are correlated across the blood-CSF barrier. Extensive mechanistic studies are needed to determine the functional impact of the metabolic changes found in this study and establish the true therapeutic potential of the metabolic changes observed post-MMKD.

## Methods

### Study participants

20 individuals were recruited who were at risk for AD based on their baseline cognitive impairment (MCI), which was diagnosed by experienced physicians and neuropsychologists using either NIA-AA MCI criteria or Alzheimer’s Disease Neuroimaging Initiative (ADNI) criteria for subjective memory complaints (SMC)^71^. All participants had prediabetes according to the American Diabetes Association guidelines, with a screening hemoglobin A1c level of 5.7-6.4%^72^. We excluded individuals who had previously been diagnosed with neurological or neurodegenerative diseases (except for MCI), major psychiatric disorders (although well-controlled depression was permitted), prior strokes, or who were currently taking diabetes or lipid-lowering medications, or medications that affect the central nervous system, such as anti-seizure drugs, antipsychotics, or opioids.

The study protocol was approved by the Wake Forest Institutional Review Board and registered with ClinicalTrials.gov (Identifier: NCT02984540). Written informed consent was obtained from all participants and their study partners, and medical professionals supervised them while the Wake Forest Institutional Data and Safety Monitoring Committee oversaw safety monitoring.

### Diet intervention procedure

The study was a pilot trial conducted in a randomized crossover design, where participants were randomly assigned to follow either the Modified Mediterranean-Ketogenic Diet (MMKD) or the American Heart Association Diet (AHAD) for a duration of 6 weeks. This was followed by a 6-week washout period, during which participants resumed their pre-study diet, before switching to the other assigned diet for another 6 weeks. The initial diet assignment was randomized using a random number generator. For more detailed information, refer to Neth et al^20^.

The experimental diet (MMKD) was a modified form of the ketogenic diet that is increasingly used to treat medically intractable epilepsy due to its better tolerability and similar effectiveness compared to a traditional ketogenic diet^73^. The targeted macronutrient composition for the MMKD was 5-10% carbohydrate, 60-65% fat, and 30% protein. Participants were instructed to consume less than 20g of carbohydrates per day and to avoid store-bought products marketed as “low carbohydrate” and artificially sweetened beverages. They were encouraged to consume extra virgin olive oil, fish, lean meats, and nutrient-rich foods such as green leafy vegetables, nuts, and berries as a source of carbohydrates.

The control diet (AHAD) was adapted from the low-fat American Heart Association Diet^74^. The target macronutrient composition for the AHAD was 55-65% carbohydrate, 15-20% fat, and 20-30% protein, with a daily fat intake target of less than 40 g/day. Participants were encouraged to consume plenty of fruits, vegetables, and fiber-rich carbohydrates.

Based on a 3-day food diary, as well as participants’ body composition and activity level, a registered dietitian developed personalized daily meal plans for each participant. Participants received weekly diet education visits either in-person or over the phone, starting one week before the start of each diet and continuing throughout the intervention.

Participants were required to maintain a daily food record that was reviewed during these visits. Both diets were designed to be isocaloric and to maintain the participant’s baseline caloric needs, with the goal of keeping their weight stable throughout the study. Participants were asked to maintain their exercise and physical activity level throughout the study. Adherence to the diets was assessed using capillary ketone body (beta-hydroxybutyrate) measurements using the Nova Max Plus® and through self-reports from the participants. Participants were given a food stipend of $25/week to defray higher food costs and were provided with a daily multivitamin supplement (Centrum® Silver®) during both diets. The use of supplements such as resveratrol, CoQ10, coconut oil, and curcumin was not allowed during the study.

### Neuropsychological evaluation

At the beginning of the study and after each diet, the participants underwent assessments of immediate and delayed memory, including the Free and Cued Selective Reminding Test (FCSRT)^75^, Story Recall (a modified version of the episodic memory measure from the Wechsler Memory Scale-Revised)^76^, and the ADAS-Cog12^77^. Cognitive abilities were evaluated before and after each diet, as well as during follow-up. To minimize the effect of learning on cognitive performance, various versions of the selected tests were used.

### Blood/CSF collection and processing

Fasting blood samples were taken from each participant before and after completing each diet and during follow-up. The samples were collected in tubes and immediately placed on ice. Within 30 minutes, the tubes were spun in a cold centrifuge at 2200 rpm for 15 minutes. After that, plasma, serum, and red blood cells were separated into different storage tubes and flash frozen at a temperature of −80°C until further analysis.

Study participants underwent a lumbar puncture to collect cerebrospinal fluid (CSF) after fasting for 12 hours at the beginning of the study and after each diet. Depending on the clinician’s preference, participants were positioned either in a seated or lateral decubitus position. To numb the area, a 25-gauge needle was used to inject 1% lidocaine locally into the L3-4 or L4-5 interspace. A 22-gauge Sprotte needle was then used to withdraw up to 25 ml of CSF into sterile polypropylene tubes. The first 3 ml of CSF was sent to a local laboratory for cell count, protein, and glucose analysis. The remaining CSF was transferred into pre-chilled polypropylene tubes in 0.2 ml aliquots, frozen immediately on dry ice, and stored at −80°C for further analysis.

### Nightingale NMR quantification

A targeted, high-throughput ^1^H-NMR metabolomics platform (Nightingale Health Ltd, Helsinki, Finland) was used to quantify 250 circulating metabolites, lipid, and lipoprotein lipid measures in serum and 28 metabolites in CSF^78^. This high-throughput metabolomics platform provides absolute concentration quantification of lipids, lipid concentrations of 14 lipoprotein subclasses and major subfractions, and further abundant fatty acids in serum, as well as amino acids, ketone bodies, and energy-related metabolites in serum and CSF. Of these measurements, 165 are directly measured metabolites and 85 are derived measurements related to lipid ratios and lipoprotein composition. All measurements are listed in Supplementary Table 2. This NMR platform is based on a standardized protocol described elsewhere^78,79^.

### Statistical analysis

#### Preprocessing serum

250 biomarkers were measured in 102 serum samples. 1 biomarker with over 40% missing values was removed from the analysis. The 77 biomarker values in percentages were central log-ratio transformed and the rest of the biomarkers were log2-transformed. The remaining missing values were imputed using the k-nearest-neighbor algorithm. 4 outlier samples were removed using the LOF method, and extreme outlying biomarker values were imputed using a kNN algorithm. There were 19 paired participants in the MMKD diet group and 18 paired participants in the AHA diet group.

#### Preprocessing CSF

28 biomarkers were measured in 59 CSF samples. None of the biomarkers or samples had more than 40% missing values. The biomarkers were log2-transformed, and the missing values were imputed using the k-nearest-neighbor algorithm. No outlier samples were detected using the LOF method, and extreme outlying biomarker values were imputed using a kNN algorithm. There were 19 paired participants in both diet groups. All preprocessing was performed with the R package maplet (https://github.com/krumsieklab/maplet)^80^.

#### Differential abundance analysis

To identify the metabolites altered due to diet, diagnosis, or the interaction of the two, a linear mixed effect model with the form *metabolite ∼ timepoint*diagnosis + age + sex + (1 | subjectID)* was used. The timepoint is a binary variable indicating measurements pre- and post-diet. The analysis was performed per diet and per fluid. P-values of the associations were adjusted using the BH method. Due to the low sample size (19) and high number of metabolites (249) tested, for serum-based analysis, a less conservative FDR threshold of 20% was used to determine significance. For CSF, the more stringent threshold of 5% was used as fewer metabolites (28) were tested. All scripts for analysis and plotting are available at https://github.com/krumsieklab/keto-beam-ad

#### Biomarker annotations

Most of the metabolites were grouped using Nightingale-provided annotations. In addition, some metabolic groups were more precisely annotated through further literature review, namely creatine and creatinine in the ATP cycle^81^ and branched-chain amino acid degradation products measured in CSF^82^.

### Metagenomic sample processing

DNA was extracted from stool samples according to Earth Microbiome Project protocols^83^ using the QIAGEN® MagAttract® PowerSoil® DNA KF Kit (384-sample). A total of 5 ng (or 3.5 µL maximum) genomic DNA was used in a 1:10 miniaturized Kapa HyperPlus protocol with a 15-cycle PCR amplification for shotgun metagenomic sequencing. Libraries were quantified with the PicoGreen dsDNA assay kit, and 50 ng (or 1 µL maximum) of each library was pooled. The pool was size selected for 300 to 700 bp and sequenced as a paired-end 150-cycle run on an Illumina HiSeq 4000 sequencer at the UCSD IGM Genomics Center^84^.

### Metagenomic data processing

Shotgun sequencing data were uploaded to and processed by Qiita^85^ (Study ID 13662). Human reads were removed using minimap2 2.17^86^, while adapters, quality filtering, and trimming were performed using fastp 20.1^87^. Remaining reads were recruited to the Web of Life database^88^ with Bowtie2 v2.3.0^89^ using the parameters from the SHOGUN pipeline^90^, then processed into Operational Genomic Units with Woltka^91^. The resulting feature table was used for downstream analysis.

### Relative abundance of features analysis

We identified metagenomic, food-omics, and metabolomic features that are associated with diet and cognition by performing Bayesian inferential regression (https://github.com/gibsramen/BIRDMAn). We utilized a Negative Binomial Linear Mixed Effect model to ensure that our statistics agreed with our study design; we modeled time, dietary sequence (whether MMKD intervention was first or second), cognitive status, and diet (whether a given individual was on MMKD or AHAD at a given time point) as fixed effects while subject identity was modeled as a random effect. Microbes, food features, and metabolites were ordered by the log-ratio of their relative abundances in objectively normal cognition to mild cognitive impairment or MMKD to AHAD individuals, respectively^92,93^ and the top and bottom ten features were examined more closely^21^.

### Multi-omics analysis

To perform a multi-omics analysis in line with the repeated-measure study design we performed joint tensor factorization. The complete methodology including mathematical formulas for joint tensor factorization can be found in Supplementary Methods. Briefly, each matrix is then transformed, through the centered-log-ratio transformation (clr) with a pseudo count of 1 to center the data around zero and approximate a normal distribution. The joint tensor factorization used here is built upon decomposition of a single temporal tensor using an approximately CP low-rank structure with multiple tensors using a shared subject matrix^94–96^. The correlations of all features across all input matrices are calculated from the final estimated matrices. The metabolic capabilities of the microbes were evaluated with MetaCyc^97^ on the Web of Life genomes^88^. Any genomes containing ilvB, ilvC, ilvD, ilvN were considered BCAA biosynthesis containing/encoding^35^.

## Data Availability

Samples were provided by Wake Forest Alzheimer's Disease Research Center. Clinical data can be requested from the National Alzheimer’s Coordinating Center.

https://naccdata.org/

## Acknowledgments and Funding

Samples were provided by Wake Forest Alzheimer’s Disease Research Center (WFADRC) supported by (P30AG049638) and associated laboratory staff, the Hartman Family Foundation, and the Roena B. Kulynych Center for Memory and Cognition Research. We would like to acknowledge the contributions of the Wake Forest Clinical and Translational Science Institute which is supported by the National Center for Advancing Translational Sciences (NCATS) through National Institutes of Health grant UL1TR001420. Additional support was provided by the Swedish Research Council and Swedish State Support for Clinical Research (ALFGBG) and laboratory technicians at Sahlgrenska University Hospital, Mölndal, Sweden. Metabolomics data is provided by the Alzheimer’s Gut Microbiome Project (AGMP) or the Alzheimer’s Disease Metabolomics Consortium (ADMC) funded wholly or in part by the following grants and supplements thereto: NIA R01AG046171, RF1AG051550, RF1AG057452, R01AG059093, RF1AG058942, U01AG061359, U19AG063744 and FNIH: #DAOU16AMPA awarded to Dr. Kaddurah-Daouk at Duke University in partnership with a large number of academic institutions. As such, the investigators within the AGMP and the ADMC, not listed specifically in this publication’s author’s list, provided data along with its pre-processing and prepared it for analysis, but did not participate in analysis or writing of this manuscript. A listing of AGMP Investigators can be found at https://alzheimergut.org/meet-the-team/. A complete listing of ADMC investigators can be found at: https://sites.duke.edu/adnimetab/team/. Drs. Krumsiek and Batra are supported by the National Institute of Aging of the National Institutes of Health under awards U19AG063744, R01AG069901-01 and Alzheimer’s association award AARFD-22-974775.

## References

1. Hampel, H. et al. Blood-based biomarkers for Alzheimer disease: mapping the road to the clinic. Nat. Rev. Neurol. 14, 639–652 (2018).

2. 2022 Alzheimer’s disease facts and figures. Alzheimer’s Dement. 18, 700–789 (2022).

3. Golde, T. E. Disease-Modifying Therapies for Alzheimer’s Disease: More Questions than Answers. Neurotherapeutics 19, 209–227 (2022).

4. Dhillon, S. Aducanumab: First Approval. Drugs 81, 1437–1443 (2021).

5. Shi, M., Chu, F., Zhu, F. & Zhu, J. Impact of Anti-amyloid-β Monoclonal Antibodies on the Pathology and Clinical Profile of Alzheimer’s Disease: A Focus on Aducanumab and Lecanemab. Frontiers in Aging Neuroscience 14, (2022).

6. Ballard, C. et al. Alzheimer’s disease. Lancet 377, 1019–1031 (2011).

7. Armstrong, R. A. Risk factors for Alzheimer’s disease. Folia Neuropathol. 57, 87–105 (2019).

8. Giri, M., Zhang, M. & Lü, Y. Genes associated with Alzheimer’s disease: an overview and current status. Clin. Interv. Aging 11, 665–681 (2016).

9. Skoog, I. & Gustafson, D. Update on hypertension and Alzheimer’s disease. Neurol. Res. 28, 605–611 (2006).

10. Kimura, N. Diabetes Mellitus Induces Alzheimer’s Disease Pathology: Histopathological Evidence from Animal Models. International Journal of Molecular Sciences 17, (2016).

11. Solomon, A. et al. Serum cholesterol changes after midlife and late-life cognition. Neurology 68, 751 LP – 756 (2007).

12. Silva, M. V. F. et al. Alzheimer’s disease: risk factors and potentially protective measures. J. Biomed. Sci. 26, 33 (2019).

13. Tabaie, E. A., Reddy, A. J. & Brahmbhatt, H. A narrative review on the effects of a ketogenic diet on patients with Alzheimer’s disease. AIMS Public Heal. 9, 185 (2022).

14. Stafstrom, C. E. & Rho, J. M. The Ketogenic Diet as a Treatment Paradigm for Diverse Neurological Disorders. Front. Pharmacol. 3, (2012).

15. Allen, B. G. et al. Ketogenic diets as an adjuvant cancer therapy: History and potential mechanism. Redox Biol. 2, 963–970 (2014).

16. Yang, L. et al. Ketogenic diet and chemotherapy combine to disrupt pancreatic cancer metabolism and growth. Med (New York, N.Y.) 3, 119–136 (2022).

17. Luukkonen, P. K. et al. Effect of a ketogenic diet on hepatic steatosis and hepatic mitochondrial metabolism in nonalcoholic fatty liver disease. Proc. Natl. Acad. Sci. 117, 7347–7354 (2020).

18. Dashti, H. M. et al. Long Term Effects of Ketogenic Diet in Obese Subjects with High Cholesterol Level. Mol. Cell. Biochem. 286, 1–9 (2006).

19. Taylor, M. K., Sullivan, D. K., Mahnken, J. D., Burns, J. M. & Swerdlow, R. H. Feasibility and efficacy data from a ketogenic diet intervention in Alzheimer’s disease. Alzheimer’s Dement. Transl. Res. Clin. Interv. 4, 28–36 (2018).

20. Neth, B. J. et al. Modified ketogenic diet is associated with improved cerebrospinal fluid biomarker profile, cerebral perfusion, and cerebral ketone body uptake in older adults at risk for Alzheimer’s disease: a pilot study. Neurobiol. Aging 86, 54–63 (2020).

21. Dilmore, A. H. et al. Effects of a ketogenic and low-fat diet on the human metabolome, microbiome, and foodome in adults at risk for Alzheimer’s disease. Alzheimer’s Dement. **n/a**, (2023).

22. Krikorian, R. et al. Dietary ketosis enhances memory in mild cognitive impairment. Neurobiol. Aging 33, 425.e19–425.e27 (2012).

23. Masino, S. A., Ruskin, D. N., Freedgood, N. R., Lindefeldt, M. & Dahlin, M. Differential ketogenic diet-induced shift in CSF lipid/carbohydrate metabolome of pediatric epilepsy patients with optimal vs. no anticonvulsant response: a pilot study. Nutr. Metab. (Lond). 18, 23 (2021).

24. Effinger, D. et al. A ketogenic diet substantially reshapes the human metabolome. Clin. Nutr. 42, 1202–1212 (2023).

25. Neth, B. J. et al. Therapeutic potential of a modified Mediterranean ketogenic diet in reversing the peripheral lipid signature of Alzheimer’s disease. medRxiv 2023.06.13.23291049 (2023). doi:10.1101/2023.06.13.23291049

26. Wallace, D. C., Fan, W. & Procaccio, V. Mitochondrial energetics and therapeutics. Annu. Rev. Pathol. 5, 297–348 (2010).

27. Liu, J. et al. Longitudinal analysis of UK Biobank participants suggests age and APOE-dependent alterations of energy metabolism in development of dementia. medRxiv 2022.02.25.22271530 (2022). doi:10.1101/2022.02.25.22271530

28. Veech, R. L. The therapeutic implications of ketone bodies: the effects of ketone bodies in pathological conditions: ketosis, ketogenic diet, redox states, insulin resistance, and mitochondrial metabolism. *Prostaglandins*, Leukot. Essent. Fat. Acids 70, 309–319 (2004).

29. Barbagallo, M. & Dominguez, L. J. Type 2 diabetes mellitus and Alzheimer’s disease. World J. Diabetes 5, 889–893 (2014).

30. Nutrition and Traumatic Brain Injury: Improving Acute and Subacute Health Outcomes in Military Personnel. (National Academies Press, 2011). doi:10.17226/13121

31. Holeček, M. Why Are Branched-Chain Amino Acids Increased in Starvation and Diabetes? Nutrients 12, (2020).

32. Krug, S. et al. The dynamic range of the human metabolome revealed by challenges. FASEB J. Off. Publ. Fed. Am. Soc. Exp. Biol. 26, 2607–2619 (2012).

33. Wheless, J. W. History of the ketogenic diet. Epilepsia 49, 3–5 (2008).

34. Toledo, J. B. et al. Metabolic network failures in Alzheimer’s disease: A biochemical road map. Alzheimers Dement 13, 965–984 (2017).

35. Zhang, Y. et al. Plasma branched-chain and aromatic amino acids correlate with the gut microbiota and severity of Parkinson’s disease. *npj Park*. Dis. 8, 48 (2022).

36. Pedersen, H. K. et al. Human gut microbes impact host serum metabolome and insulin sensitivity. Nature 535, 376–381 (2016).

37. Chiesa, S. et al. Glycoprotein Acetyls: A Novel Inflammatory Biomarker of Early Cardiovascular Risk in the Young. J. Am. Heart Assoc. 11, (2022).

38. Ballout, R. A. & Remaley, A. T. GlycA: A New Biomarker for Systemic Inflammation and Cardiovascular Disease (CVD) Risk Assessment. J. Lab. Precis. Med. 5, (2020).

39. Calice-Silva, V., Suemoto, C. K., Brunoni, A. R., Bensenor, I. M. & Lotufo, P. A. Association Between GlycA and Cognitive Function: Cross-Sectional Results From the ELSA—Brasil Study. Alzheimer Dis. Assoc. Disord. 35, (2021).

40. Marsland, A. L. et al. Interleukin-6 covaries inversely with cognitive performance among middle-aged community volunteers. Psychosom. Med. 68, 895–903 (2006).

41. Holmes, C. et al. Systemic inflammation and disease progression in Alzheimer disease. Neurology 73, 768–774 (2009).

42. Pinto, A., Bonucci, A., Maggi, E., Corsi, M. & Businaro, R. Anti-Oxidant and Anti-Inflammatory Activity of Ketogenic Diet: New Perspectives for Neuroprotection in Alzheimer’s Disease. *Antioxidants (Basel*, Switzerland) 7, (2018).

43. Dupuis, N., Curatolo, N., Benoist, J.-F. & Auvin, S. Ketogenic diet exhibits anti-inflammatory properties. Epilepsia 56, e95–8 (2015).

44. Ouimet, M., Barrett, T. J. & Fisher, E. A. HDL and Reverse Cholesterol Transport. Circ. Res. 124, 1505–1518 (2019).

45. Svensson, T. et al. The association between midlife serum high-density lipoprotein and mild cognitive impairment and dementia after 19 years of follow-up. Transl. Psychiatry 9, 26 (2019).

46. Batra, R. et al. The landscape of metabolic brain alterations in Alzheimer’s disease. Alzheimer’s Dement. 19, 980–998 (2023).

47. Chuang, D. T., Chuang, J. L. & Wynn, R. M. Lessons from Genetic Disorders of Branched-Chain Amino Acid Metabolism1, 2, 3. J. Nutr. 136, 243S–249S (2006).

48. Kanehisa, M., Sato, Y., Kawashima, M., Furumichi, M. & Tanabe, M. KEGG as a reference resource for gene and protein annotation. Nucleic Acids Res. 44, D457–D462 (2016).

49. Biswas, D., Duffley, L. & Pulinilkunnil, T. Role of branched-chain amino acid–catabolizing enzymes in intertissue signaling, metabolic remodeling, and energy homeostasis. FASEB J. 33, 8711–8731 (2019).

50. Redjems-Bennani, N. et al. Abnormal substrate levels that depend upon mitochondrial function in cerebrospinal fluid from Alzheimer patients. Gerontology 44, 300–304 (1998).

51. van der Velpen, V. et al. Systemic and central nervous system metabolic alterations in Alzheimer’s disease. Alzheimers. Res. Ther. 11, 93 (2019).

52. Ibáñez, C. et al. A new metabolomic workflow for early detection of Alzheimer’s disease. J. Chromatogr. A 1302, 65–71 (2013).

53. Ibáñez, C. et al. Toward a predictive model of Alzheimer’s disease progression using capillary electrophoresis-mass spectrometry metabolomics. Anal. Chem. 84, 8532–8540 (2012).

54. Berezhnoy, G., Laske, C. & Trautwein, C. Metabolomic profiling of CSF and blood serum elucidates general and sex-specific patterns for mild cognitive impairment and Alzheimer’s disease patients. Front. Aging Neurosci. 15, 1219718 (2023).

55. Ghersi-Egea, J.-F. et al. Molecular anatomy and functions of the choroidal blood-cerebrospinal fluid barrier in health and disease. Acta Neuropathol. 135, 337–361 (2018).

56. Felby, S., Nielsen, E. & Thomsen, J. L. The postmortem distribution of ketone bodies between blood, vitreous humor, spinal fluid, and urine. Forensic Sci. Med. Pathol. 4, 100– 107 (2008).

57. Achanta, L. B. & Rae, C. D. β-Hydroxybutyrate in the Brain: One Molecule, Multiple Mechanisms. Neurochem. Res. 42, 35–49 (2017).

58. Newman, J. C. & Verdin, E. Ketone bodies as signaling metabolites. Trends Endocrinol. Metab. 25, 42–52 (2014).

59. Chu, C. Q. et al. Can dietary patterns prevent cognitive impairment and reduce Alzheimer’s disease risk: Exploring the underlying mechanisms of effects. Neurosci. Biobehav. Rev. 135, 104556 (2022).

60. Lilamand, M. et al. Are ketogenic diets promising for Alzheimer’s disease? A translational review. Alzheimer’s Res. Ther. 12, 1–10 (2020).

61. Marsland, A. L., Gianaros, P. J., Abramowitch, S. M., Manuck, S. B. & Hariri, A. R. Interleukin-6 covaries inversely with hippocampal grey matter volume in middle-aged adults. Biol. Psychiatry 64, 484–490 (2008).

62. Toledo, J. B. et al. Metabolic network failures in Alzheimer’s disease: A biochemical road map. Alzheimers. Dement. 13, 965–984 (2017).

63. Neth, B. J. et al. Therapeutic potential of a modified Mediterranean ketogenic diet in reversing the peripheral lipid signature of Alzheimer’s disease. medRxiv 2023<otherinfo>.06.13.23291049 (2023). doi:10.1101/2023.06.13.23291049</otherinfo>

64. Newman, A. B. et al. Dementia and Alzheimer’s Disease Incidence in Relationship to Cardiovascular Disease in the Cardiovascular Health Study Cohort. J. Am. Geriatr. Soc. 53, 1101–1107 (2005).

65. Pedditzi, E., Peters, R. & Beckett, N. The risk of overweight/obesity in mid-life and late life for the development of dementia: a systematic review and meta-analysis of longitudinal studies. Age Ageing 45, 14–21 (2016).

66. Anstey, K. J., Cherbuin, N., Budge, M. & Young, J. Body mass index in midlife and late-life as a risk factor for dementia: a meta-analysis of prospective studies. Obes. Rev. an Off. J. Int. Assoc. Study Obes. 12, e426–37 (2011).

67. Butterfield, D. A. & Halliwell, B. Oxidative stress, dysfunctional glucose metabolism and Alzheimer disease. Nat. Rev. Neurosci. 20, 148–160 (2019).

68. Roy, M., et al. A ketogenic supplement improves white matter energy supply and processing speed in mild cognitive impairment. Alzheimer’s Dement. (New York, N. Y.) 7, e12217 (2021).

69. Roy, M. et al. A ketogenic intervention improves dorsal attention network functional and structural connectivity in mild cognitive impairment. Neurobiol. Aging 115, 77–87 (2022).

70. Safieh, M., Korczyn, A. D. & Michaelson, D. M. ApoE4: an emerging therapeutic target for Alzheimer’s disease. BMC Med. 17, 64 (2019).

71. Risacher, S. L. et al. APOE effect on Alzheimer’s disease biomarkers in older adults with significant memory concern. Alzheimers. Dement. 11, 1417–1429 (2015).

72. Standards of medical care in diabetes--2011. Diabetes Care 34 Suppl 1, (2011).

73. Kossoff, E. H. & Dorward, J. L. The modified Atkins diet. Epilepsia 49 **Suppl 8**, 37–41 (2008).

74. Krauss, R. M. et al. AHA Dietary Guidelines. Circulation 102, 2284–2299 (2000).

75. Grober, E., Sanders, A. E., Hall, C. & Lipton, R. B. Free and cued selective reminding identifies very mild dementia in primary care. Alzheimer Dis. Assoc. Disord. 24, 284–290 (2010).

76. Wechsler, D. Wechsler Memory Scale-Revised. (Psychological Corporation, 1987).

77. Rosen, W. G., Mohs, R. C. & Davis, K. L. A new rating scale for Alzheimer’s disease. Am. J. Psychiatry 141, (1984).

78. Soininen, P., Kangas, A. J., Würtz, P., Suna, T. & Ala-Korpela, M. Quantitative serum nuclear magnetic resonance metabolomics in cardiovascular epidemiology and genetics. Circ. Cardiovasc. Genet. 8, 192–206 (2015).

79. Soininen, P. et al. High-throughput serum NMR metabonomics for cost-effective holistic studies on systemic metabolism. Analyst 134, 1781–1785 (2009).

80. Chetnik, K. et al. maplet: an extensible R toolbox for modular and reproducible metabolomics pipelines. Bioinformatics 38, 1168–1170 (2022).

81. Wyss, M. & Kaddurah-Daouk, R. Creatine and Creatinine Metabolism. Physiol. Rev. 80, 1107–1213 (2000).

82. Liebich, H. M. & Först, C. Hydroxycarboxylic and oxocarboxylic acids in urine:products from branched-chain amino acid degradation and from ketogenesis. J. Chromatogr. B Biomed. Sci. Appl. 309, 225–242 (1984).

83. Marotz, C. et al. DNA extraction for streamlined metagenomics of diverse environmental samples. Biotechniques 62, 290–293 (2017).

84. Marotz, C. et al. Evaluation of the Effect of Storage Methods on Fecal, Saliva, and Skin Microbiome Composition. mSystems 6, (2021).

85. Gonzalez, A. et al. Qiita: rapid, web-enabled microbiome meta-analysis. Nat. Methods 15, 796–798 (2018).

86. Li, H. Minimap2: pairwise alignment for nucleotide sequences. Bioinformatics 34, 3094– 3100 (2018).

87. Chen, S., Zhou, Y., Chen, Y. & Gu, J. fastp: an ultra-fast all-in-one FASTQ preprocessor. Bioinformatics 34, i884–i890 (2018).

88. Zhu, Q. et al. Phylogenomics of 10,575 genomes reveals evolutionary proximity between domains Bacteria and Archaea. Nat. Commun. 10, (2019).

89. Langmead, B. & Salzberg, S. L. Fast gapped-read alignment with Bowtie 2. Nat. Methods 9, 357–359 (2012).

90. Hillmann, B. et al. SHOGUN: a modular, accurate and scalable framework for microbiome quantification. Bioinformatics 36, 4088–4090 (2020).

91. Zhu, Q., et al. Phylogeny-Aware Analysis of Metagenome Community Ecology Based on Matched Reference Genomes while Bypassing Taxonomy. mSystems 7, (2022).

92. Morton, J. T. et al. Establishing microbial composition measurement standards with reference frames. Nat. Commun. 10, (2019).

93. Rahman, G. et al. BIRDMAn: A Bayesian differential abundance framework that enables robust inference of host-microbe associations. bioRxiv Prepr. Serv. Biol. (2023). doi:10.1101/2023.01.30.526328

94. Martino, C. et al. Context-aware dimensionality reduction deconvolutes gut microbial community dynamics. Nat. Biotechnol. 39, 165–168 (2021).

95. Han, R., Shi, P. & Zhang, A. R. Guaranteed Functional Tensor Singular Value Decomposition. J. Am. Stat. Assoc. (2023). doi:10.1080/01621459.2022.2153689

96. Shi, P. et al. Time-Informed Dimensionality Reduction for Longitudinal Microbiome Studies. bioRxiv 2023.07.26.550749 (2023). doi:10.1101/2023.07.26.550749

97. Caspi, R. et al. The MetaCyc database of metabolic pathways and enzymes and the BioCyc collection of Pathway/Genome Databases. Nucleic Acids Res. 42, (2014).

