## Supplemental Methods for "A Modified Mediterranean Ketogenic Diet mitigates modifiable risk factors of Alzheimer’s Disease: a serum and CSF-based metabolic analysis"

#### Joint Tensor Factorization algorithm

We adopt the model setting proposed in [1], which can be used to reduce the dimensions jointly between multiple temporal tensors formed by $\bar{T}_1$ and $\bar{T}_2$ through the objective function:

$$\min_{\lambda, a_{i,2}\xi} \sum_{i,j,t\in T_i} \left( \left(\bar{T}_1\right)_{ijt} - \lambda_1 a_i b_{ij} \xi_{it} \right)^2 + \sum_{i,j,t\in T_i} \left( \left(\bar{T}_2\right)_{ijt} - \lambda_2 a_{i,} b_{2j} \xi_2 t \right)^2$$

Where $q_1 \lambda_1, \lambda_2$ are updated separately.

$$\hat{a}_i^{(t+1)} = \underset{a}{\operatorname{argmin}} \left[ \sum_{j,t\in T_i} \left( b_{1j}^{(t)} \xi_1^{(t)} \lambda_1^{(t)} \right)^{(t)} + \left( b_{2j}^{(t)} \xi_{2t}^{(t)} \lambda_2^{(t)} \right)^2 \right\} a_i^2$$

$$\text{a} \; -2 \sum_{j,t\in T_i} \left\{ \left(\bar{T}_1\right)_{ijt} \lambda_1^{(t)} b_{1j}^{(t)} \xi_{1t}^{(t)} + \left(\bar{T}_2\right)_{ijt} \lambda_2^{(t)} b_2 j \xi_2^{(t)} s_2^{(t)} \right\} a \Bigg]$$

$$= \frac{\sum_{j\cdot t\epsilon T_i} \left\{ (\bar{T})_{ij\tau} b_{1j}^{(t)} \xi_{1t}^{(t)} \lambda_1^{(t)} + \left(\bar{T}_2\right)_{ijt} b_{2j}^{(t)} \xi_{2\tau}^{(\tau)} \lambda_2^{(t)} \right\}}{\lambda_1^{(t)} \sum_{t\in T_i} \xi_{1t}^{(t)2} + \lambda_2^{(t)2} \sum_{t\in T_i} \xi_{2t}^{(t)2}}$$

$$\hat{\lambda}_1^{(t+1)} = \arg \lambda_1 \min \left[ \sum_{i,j,t\in T_i} \lambda_1^2 \left(a_i b_{1j} \xi_1 t\right)^2 - 2 \sum_{i,j\cdot t\in T_i} \left(\bar{T}_1\right)_{ijt} a_i b_{1j} \xi_1 \lambda_1 \right]$$

$$= \frac{\sum_{i,t\in T_i} \left(\bar{T}_1\right)_{itt} a_{vi} b_{ij} \xi_{1t}}{\sum_{i,t\in T_i} \left(a_i \xi_{1t}\right)^2}$$

$$\hat{\lambda}_2^{(t+1)} = \frac{\sum_{i,j=t\in T_i} \left(\bar{T}_2\right)_{ijt} a_i b_{2j} \xi_{2t}}{\sum_{i,j\in T_i} \left(a_i \xi_{2t}\right)^2}$$

$$\hat{b}_{(j)}^{(t+1)} = \frac{\sum_{i,t\in T_i} a_i \xi_{1t} \left(\bar{T}_1\right)_{jit}}{\sum_{i,t\in \pi} \left(a_i \xi_1 t\right)^2}, \hat{b}_1^{(t+1)} = \hat{b}_1^{(t+1)} / \left\| \hat{b}^{(t+1)} \right\|_2$$

$$\hat{b}_{2j}^{(t+1)} = \frac{\sum_{i\in I_i} a_i \xi_{2t} \left(\bar{T}_2\right)_{ijt}}{\sum_{i,t\in T_i} \left(a_i \xi_{it}\right)^2}, \hat{b}_2^{(t+1)} = \hat{b}_2^{(t+1)} / \left\| \hat{b}_2^{(t+1)} \right\|_2$$

where $a$ are subject loadings, $b$ are feature loadings that can be used to identify features contributing to the beta analysis, $\xi^{(l)}(t)$ is the temporal loading that captures the shared temporal patterns among subjects and features, and $\lambda$ quantifies the contribution of each component. For each $b^1$ and $b^2$ we define $W^1$ and $W^2$ respectively given by:

$$\mathrm{W}^1 = (\lambda_1 b^{1^T})^T$$
$$\mathrm{W}^2 = (\lambda_2 b^{2^T})^T$$

The temporal covariance and correlations of all features across all input matrices are calculated from the final estimated matrices by:

$$\textit{temporal feature covariance matrix} = \begin{bmatrix} \boldsymbol{W_1} \\ \boldsymbol{W_2} \end{bmatrix} \begin{bmatrix} \boldsymbol{W_1} \\ \boldsymbol{W_2} \end{bmatrix}^T$$

Which provides the estimated relationships between the features across both input temporal tensors across time.

### References


1. Shi, P. *et al. Time-Informed Dimensionality Reduction for Longitudinal Microbiome Studies* en. July 2023. https://www.biorxiv.org/content/10.1101/2023.07.26.550749v1.
